# Potential interruptions in HIV prevention and treatment services for gay, bisexual, and other men who have sex with men associated with COVID-19

**DOI:** 10.1101/2020.08.19.20178285

**Authors:** Amrita Rao, Katherine Rucinski, Brooke Jarrett, Benjamin Ackerman, Sara Wallach, Julia Marcus, Tyler Adamson, Alex Garner, Glenn-Milo Santos, Chris Beyrer, Sean Howell, Stefan Baral

## Abstract

**Background:** Globally, the coronavirus pandemic has necessitated a range of population-based measures in order to stem the spread of infection and reduce COVID-19-related morbidity and mortality. These measures may be associated with disruptions to other health services including for gay, bisexual, and other men who have sex with men (MSM) at risk for or living with HIV. Here, we assess the relationship between stringency of COVID-19 mitigation strategies and interruptions to HIV prevention and treatment services for MSM.

**Methods:** Data for this study were collected as part of a COVID-19 Disparities Survey implemented by the gay social networking app *Hornet*, with data collected between April 16^th^, 2020 and May 24^th^, 2020. Data were assessed for countries where at least 50 participants completed the survey, to best evaluate country-level heterogeneity. We used a modified Poisson regression model, with clustering at the country-level, to assess the association between stringency of pandemic control measures and access to HIV services. Pandemic control measures were quantified using the Oxford Government Response Tracker Stringency Index; each country received a score (0-100) based on the number and strictness of nine indicators related to school and workplace closures and travel bans.

**Results:** A total of 10,654 MSM across 20 countries were included in these analyses. The mean age was 34.2 (standard deviation: 10.8), and 12% (1264/10540) of participants reported living with HIV. The median stringency score was 82.31 (Range:[19.44, Belarus]-[92.59, Ukraine]). For every ten-point increase in stringency, there was a 3% reduction in the prevalence of access to in-person testing (aPR: 0.97, 95% CI:[0.96, 0.98]), a 6% reduction in the prevalence of access to self-testing (aPR: 0.94, 95% CI:[0.93, 0.95]), and a 5% reduction in access to PrEP (aPR: 0.95, 95% CI:[0.95, 0.97]). Among those living with HIV, close to one in five (n = 218/1105) participants reported being unable to access their provider either in-person or via telemedicine during the COVID-19 pandemic, with a greater proportion of interruptions to treatment services reported in Belarus and Mexico. Almost half (n = 820/1254) reported being unable to refill their HIV medicine prescription remotely.

**Conclusions:** More stringent government responses were associated with decreased access to HIV diagnostic, prevention, and treatment services. To minimize increases in HIV-related morbidity and mortality, innovative strategies are needed to facilitate minimize service interruptions to MSM communities during this and potential future waves of COVID-19.

## INTRODUCTION

As of August, 2020, the severe acute respiratory syndrome coronavirus 2 (SARS-CoV-2) has infected more than 18 million people and resulted in more than half a million deaths worldwide.^1^ Large scale government mitigation strategies, including stay-at-home orders, restrictions on social gatherings, and the closure of public transportation, have swept the globe in an effort to control the pandemic.^2,3^ However, the scope, stringency, and timing of these mitigation strategies have varied across countries — some have implemented severe, nationwide measures, while others have implemented a more adaptive response strategy.^3^

Concurrently, the human immunodeficiency virus (HIV) pandemic newly infected 1.7 million people in 2019, for a total of 38 million living with HIV in 2020.^4^ Unlike SARS-CoV-2, lifesaving antiretroviral therapies (ART) have prevented 12.1 million AIDS-related deaths over the past ten years.^5^ Prevention strategies and treatment strategies, like pre-exposure prophylaxis for HIV prevention (PrEP) and treatment for people living with HIV, can effectively eliminate transmission risks.^6^–^9^ Despite this, barriers to HIV prevention and treatment services persist, particularly for those at highest risk of HIV acquisition. Condom use is on the decline or has leveled off as measured by condom sales, 19% of people living with HIV, or 7.1 million people, are unaware of their status, and about 33% of people living with HIV, or 12.6 million people, are not accessing ART.^5^

Gay, bisexual, and other men who have sex with men (MSM) are especially vulnerable to HIV infection and associated morbidities and mortality.^10^–^12^ Although MSM represent between 2–5% of cisgender men globally, unmet HIV prevention and treatment needs resulted in MSM accounting for nearly a quarter (24%) of new HIV infections in 2019.^5^ MSM also face barriers to prevention services and treatment specific to their sexuality, including anti-homosexuality laws as well as intersecting stigmas from healthcare workers.^13^–^17^ The SARS-CoV-2 pandemic has the potential to further exacerbate these disparities in HIV prevention and treatment.^18^

Though the 90-90-90 targets for 2020 set by the Joint United Nations Programme on HIV and AIDS were far from being met, the diversion of healthcare resources from HIV and toward SARS-CoV-2 may further dampen progress toward these targets or even reverse gains over the last several years.^19^ For instance, the World Health Organization recently reported that 73 countries are in danger of stocking out their supplies of ART, and other parts of the HIV care continuum, including access to testing and sustained viral suppression, are also threatened.^20,21^ Currently, there are limited empiric assessments of how mitigation strategies designed to slow the spread of COVID-19 will impact engagement in HIV prevention and treatment services among MSM. The objective of these analyses was to assess the relationship between the stringency of COVID-19 mitigation strategies and interruptions to HIV prevention and treatment services among MSM globally.

## METHODS

### Social Media Based Sample

Data for this study primarily came from a cross-sectional survey implemented by the gay social networking app Hornet. Hornet is a free mobile app with over 25 million global users and four million active daily users. Hornet’s primary mission is to empower and connect gay men to a larger social network and community. Between April 16, 2020 and May 24, 2020, active Hornet users were invited through their Hornet-specific inbox to participate in a survey comprising a series of questions related to the impact of COVID-19 on a range of economic and health indicators.^22^

Hornet users were eligible for the study if they had been active users of the app for at least one year and were at least 18 years old. A total of 13,563 users provided informed consent and responded to the survey, and 12,210 completed the survey. The larger study was reviewed by the Johns Hopkins School of Public Health Institutional Review Board and received a Category 4 exemption.

For this study, countries were included in analyses if they had had 50 or more individual-level responses. A total of 20 countries met this criterion: Australia, Belarus, Belgium, Brazil, Canada, Egypt, France, Germany, Indonesia, Italy, Kazakhstan, Malaysia, Mexico, the Russian Federation, Taiwan, Thailand, Turkey, Ukraine, the United Kingdom, and the United States.

### Primary Outcome Variables

Individuals were asked a series of questions related to the effects of COVID-19 on perceived access to HIV prevention and treatment services. Questions that assessed potential interruptions to HIV prevention were asked of all participants, and included access to 1) in-person HIV testing, 2) HIV self-testing, 3)PrEP, and 4) condoms. All questions began with the prompt, “Whether you need these or not, do you feel you have access to HIV prevention strategies during the COVID-19 crisis…”. Response options were based on a five-point Likert-scale: “Definitely yes,” “Probably yes,” “Might or might not,” “Probably not”, and “Definitely not”. Responses were recoded and dichotomized (“Definitely yes” and “probably yes” vs. “might or might not”, “probably not” and “definitely not”) to get a sense of overall positive or negative sentiments regarding access to each service. Questions that assessed access to HIV treatment services were asked only of those participants who reported living with HIV. These questions included 1) “Since the beginning of social-isolation, are you able to see your HIV provider if you needed to?” and 2) “Can you refill your prescription remotely?”.

### The Oxford COVID-19 Government Response Tracker

In addition to the social media-based survey, data on the stringency of COVID-19 mitigation strategies were obtained from the Oxford COVID-19 Government Response Tracker (OxCGRT).^3^ The OxCGRT tool collects data about government responses to COVID-19 and collates that data into 17 indicators: eight on containment and closures (e.g., school closing, stay-at-home requirements, and travel restrictions), four on economic policies (e.g. fiscal measures, income support, etc.), and five on health system policies (e.g., public information campaigns, contact tracing, etc.).^3^ These indicators are then aggregated to create different indices that measure government action. In this study, we used the OxCGRT “Government Response Stringency Index.”^3^

### Primary Exposure Variable

The OxCGRT Government Response Stringency Index is based on nine of the 17 available indicators and includes all indicators related to containment and closures and one indicator of health system policies.^3^ Specifically, the containment and closures indicators reflect school and workplace closures, public events cancellations and restrictions on gatherings, closure of public transportation, stay-at-home requirements and restrictions on internal movement, and international travel controls.^3^ The health system indicator relates to whether public information campaigns related to COVID-19 are in place (e.g. urging caution about risk of COVID-19 and coordinated mitigation efforts).^3^ A score for each indicator is created by taking the ordinal value for that indicator, adding an extra 0.5 point if the policy is “general” (i.e. applying across a jurisdiction) rather than “targeted” (i.e. applying primarily to a specific locality). Each indicator score is then rescaled by its maximum value and aggregated to create a stringency score between 0 and 100.^3^ For this study, each included country was matched to its specific stringency score as of April 6^th^, ensuring the exposure assessment preceded the launch of the app-based survey on April 16^th^. In sensitivity analyses, stringency scores were re-matched using updated assessments from April 16^th^, May 4^th^, and May 24^th^ to ensure that results were robust to any within-country changes in the stringency of responses over time **(Appendix)**. A full description of the tracker, the indicators, and the different indices to measure government action are publicly available through the *Oxford COVID-19 Government Response Tracker”* online reporting systems (https://www.bsg.ox.ac.uk/research/research-proiects/coronavirus-government-response-tracker).^3^

### Statistical Analyses

Characteristics of participants were compared by country, using one-way ANOVAs to test differences between means for continuous variables and Fisher’s exact tests to test differences in proportions for categorical variables. Mixed effects generalized linear models with modified Poisson regression, robust variance estimation, and clustering by country were used to examine the associations between stringency of government response to COVID-19 and access to HIV prevention services.^23^ Country-level Human Development Index^24^ and total health spending per capita^25^ were included for adjustment in multivariable analyses. Country-level covariates were selected for inclusion in the final model, and not individual-level variables, as they may represent common causes of both the exposure and the outcome. To aid in the interpretation of regression coefficients, the primary exposure (stringency of government response) was scaled by a factor of 10. Multi-level modeling was not performed for the HIV treatment access outcomes, given the few individuals living with HIV per included country.

### Missing Data

Missingness of all primary outcome variables was examined. Approximately 2% were missing responses for in-person testing (n = 258), 12% for self-testing (n = 1319), 14% for PrEP (n = 1481), and 10% for condoms (n = 1112). About 13% of those living with HIV were missing data on whether or not they were able to access their provider (159/1264), and 0.7% were missing data on their access to remote refills (10/1264). Based on an examination of the correlation matrix between the primary outcomes and other available data, missingness did not follow any observable pattern, and therefore complete case analysis for each of the regression analyses was utilized.

## RESULTS

### Individual-Level Characteristics

A total of 13,563 individuals initiated the survey, of whom 12,210 completed the full questionnaire. Of these respondents, 10,654 came from one of the 20 countries with more than 50 individuals represented in the dataset and were thus included in these analyses. Approximately one third of included individuals were from the Russian Federation (n = 3436), one fifth from Turkey (n = 2280), and nine percent or less from each of the remaining countries. The mean age of participants was 34.2 years (standard deviation = ll), with significantly older mean age among included individuals from Australia and the United States and younger mean age among included individuals from Belarus and Kazakhstan. About 12% (n = 1264) reported living with HIV, with a greater proportion seen among individuals from Brazil and Mexico (25.5% and 29.1%, respectively). Overall, 49% reported being of lower (n = 937) or lower middle class (n = 4254), and 15% (n = 1599) reported having no health insurance. **(Table 1)**

**Table 1.** Characteristics of gay and other men who have sex with men participants by country from a global survey implemented via the social media app *Hornet* between April 16^th^ and May 24^th^, 2020.

|  | Proportion of overall sample (n) | Human Development Index (HDI) | Total health spending per capita (USD) | Mean age (standard deviation)<sup>a</sup> | Proportion living with HIV (n)<sup>b, c, f</sup> | Proportion “lower” socioeconomic status (n)<sup>b, d, g</sup> | Proportion without insurance (n)<sup>b, e</sup> |
| --- | --- | --- | --- | --- | --- | --- | --- |
| **OVERALL** | 100% (10654) |  |  | 34.2 (10.8) | 12.0% (1264) | 8.9% (937) | 15.2% (1599) |
| **Australia** | 0.5% (54) | 0.938 | 4400 | 47.6 (13.5) | 11.5% (6) | 3.8% (2) | 7.7% (4) |
| **Belarus** | 2.2% (235) | 0.817 | 1232 | 30.1 (9.0) | 5.6% (13) | 2.6% (6) | 23.3% (54) |
| **Belgium** | 0.5% (58) | 0.919 | 4939 | 44.0 (11.0) | 17.5% (10) | 1.8% (1) | 1.8% (1) |
| **Brazil** | 6.1% (653) | 0.761 | 1431 | 38.5 (12.0) | 25.5% (166) | 11.4% (74) | 21.0% (136) |
| **Canada** | 0.6% (68) | 0.922 | 4921 | 45.6 (13.7) | 7.4% (5) | 2.9% (2) | 1.5% (1) |
| **Egypt** | 0.8% (80) | 0.700 | 484 | 33.4 (7.2) | 10.8% (8) | 11.7% (9) | 45.3% (34) |
| **France** | 9.0% (962) | 0.891 | 4741 | 42.0 (11.5) | 10.5% (100) | 3.5% (33) | 1.8% (17) |
| **Germany** | 0.7% (75) | 0.939 | 5532 | 41.8 (11.5) | 11.0% (8) | 9.5% (7) | 6.8% (5) |
| **Indonesia** | 3.9% (420) | 0.707 | 383 | 31.5 (8.3) | 11.6% (47) | 18.3 (76) | 29.5% (119) |
| **Italy** | 0.8% (81) | 0.883 | 3445 | 44.2 (11.1) | 7.4% (6) | 6.3% (5) | 14.8% (12) |
| **Kazakhstan** | 1.4% (148) | 0.817 | 1017 | 30.9 (8.2) | 10.1% (15) | 4.7% (7) | 31.5% (46) |
| **Malaysia** | 0.5% (55) | 0.804 | 1072 | 35.4 (7.9) | 20.0% (11) | 9.1% (5) | 16.4% (9) |
| **Mexico** | 2.3% (241) | 0.767 | 1081 | 36.9 (10.9) | 29.1% (70) | 4.2% (10) | 13.3% (32) |
| **Russian Federation** | 32.2% (3436) | 0.824 | 1544 | 31.4 (8.7) | 11.8% (404) | 6.5% (224) | 10.2% (349) |
| **Taiwan** | 3.1% (329) | 0.880 | 2535 | 31.5 (8.5) | 6.4% (21) | 6.7% (22) | 1.5% (5) |
| **Thailand** | 5.6% (593) | 0.765 | 614 | 35.7 (11.1) | 14.1% (83) | 13.4% (79) | 24.4% (143) |
| **Turkey** | 21.4% (2280) | 0.806 | 1029 | 32.1 (9.8) | 8.3% (184) | 13.2% (295) | 16.0% (356) |
| **Ukraine** | 5.0% (528) | 0.750 | 598 | 31.3 (9.4) | 11.5% (60) | 9.5% (50) | 47.8% (250) |
| **United Kingdom** | 2.1% (218) | 0.920 | 4285 | 44.0 (13.0) | 11.0% (23) | 5.1% (11) | 3.7% (8) |
| **United States** | 1.3% (140) | 0.920 | 9839 | 46.3 (13.3) | 17.3% (24) | 13.6% (19) | 12.9% (18) |

<sup>a</sup> Statistically significant differences between means were assessed using one-way ANOVA for continuous variables. All were significantly different at the $p < 0.01$ level.; <sup>b</sup> Statistically significant differences between proportions were assessed using Fisher’s exact tests for categorical variables. All were significantly different at the $p < 0.01$ level; <sup>c</sup> 114 individuals chose not to respond or did not know their HIV status (1.1%). <sup>d</sup> 79 individuals chose not to respond to the question asking about socioeconomic status (0.7%). <sup>e</sup> 142 individuals chose not to respond to the question asking about health insurance (1.3%). <sup>f</sup> Participants were asked “What is your HIV status? And asked to select “I’m HIV positive and undetectable,” “I’m HIV positive,” “I’m HIV negative,” “I don’t know,” or “I don’t want to answer.” <sup>g</sup> Participants were directly asked “What is your socioeconomic status?”, and asked to select “Lower,” “Lower Middle,” “Upper Middle,” or “Upper” class. <sup>h</sup> Participants were asked “Do you have health insurance?”, and asked to select “No insurance,” “Government Insurance,” or “Private/Employer/Other Non-governmental insurance.”

### Country-Level Covariates

The Human Development Index ranged in value for included countries from 0.700 in Egypt to 0.939 in Germany, compared with the full range globally of 0.377 in Niger to 0.954 in Norway.^24^ Total health spending per capita was lowest in Indonesia at 383 USD and highest in the United States of America at 9839 USD.^25^ **(Table 1)**

### Stringency of Government Responses

Stringency of government response scores ranged in value from 19.44 for Belarus to 92.59 for Ukraine. OxCGRT Government Response Stringency Index scores for each included country are reported for April 6^th^, April 16^th^, May 4^th^, and May 24^th^ in **Figure 1**.

**Figure 1.**
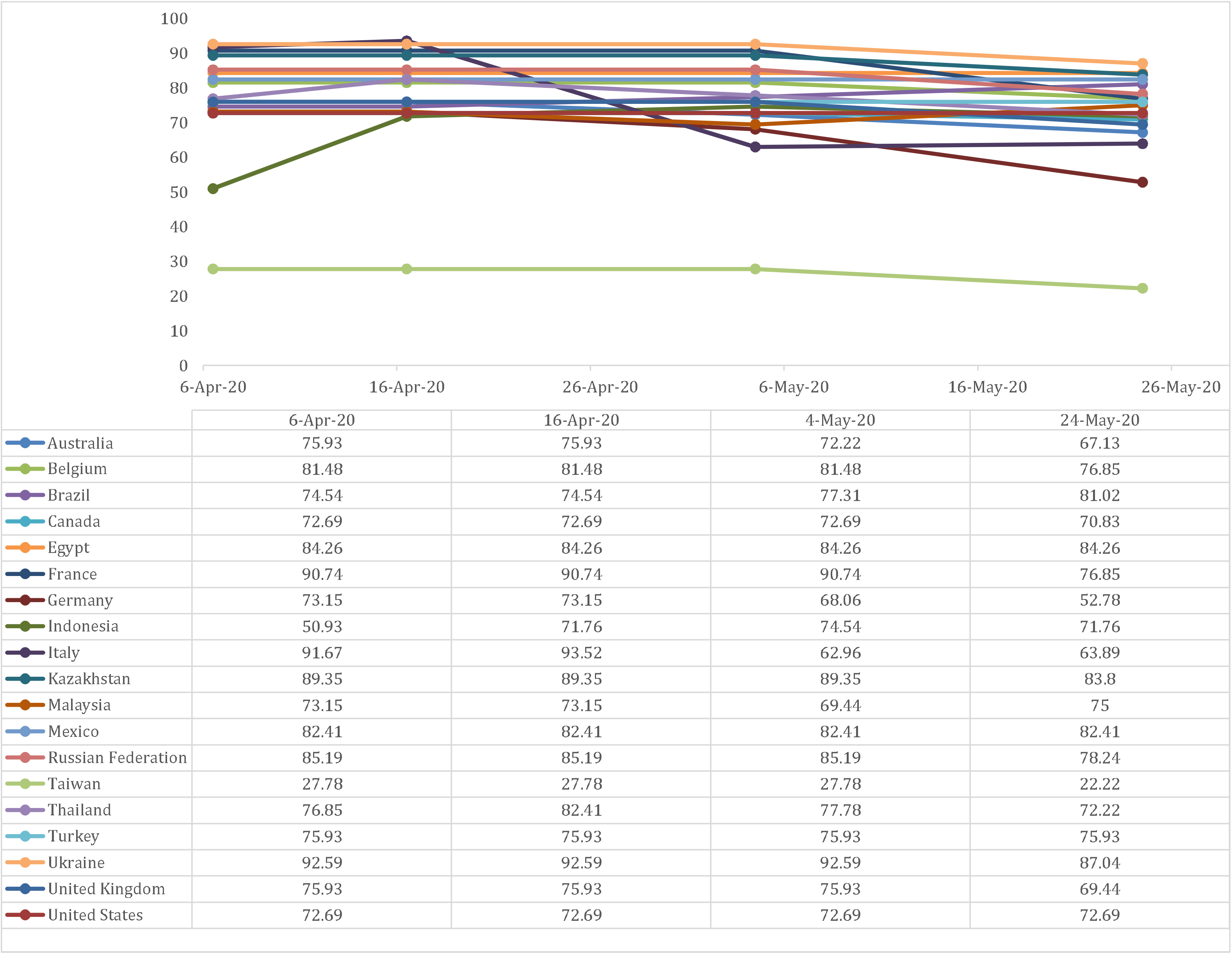
The Oxford COVID-19 Government Response Tracker (OxCGRT) Government Response Stringency Index Scores for included countries

### Access to HIV Prevention Services

Overall, 38% reported potential interruptions to in-person testing (3992/10396), with greater proportions reporting potential interruptions in Turkey and the United Kingdom. Fifty five percent (5178/9335) reported potential interruptions to HIV self-testing, 56% (5171/9173) reported potential interruptions to PrEP, and 10% (990/9542) reported potential interruptions to condom access. For HIV self-testing, greater proportions reporting potential interruptions were seen in Australia and Malaysia; for PrEP, greater proportions reporting potential interruptions were seen in Mexico and Turkey compared to all other countries **(Figure 2)**.

**Figure 2.**
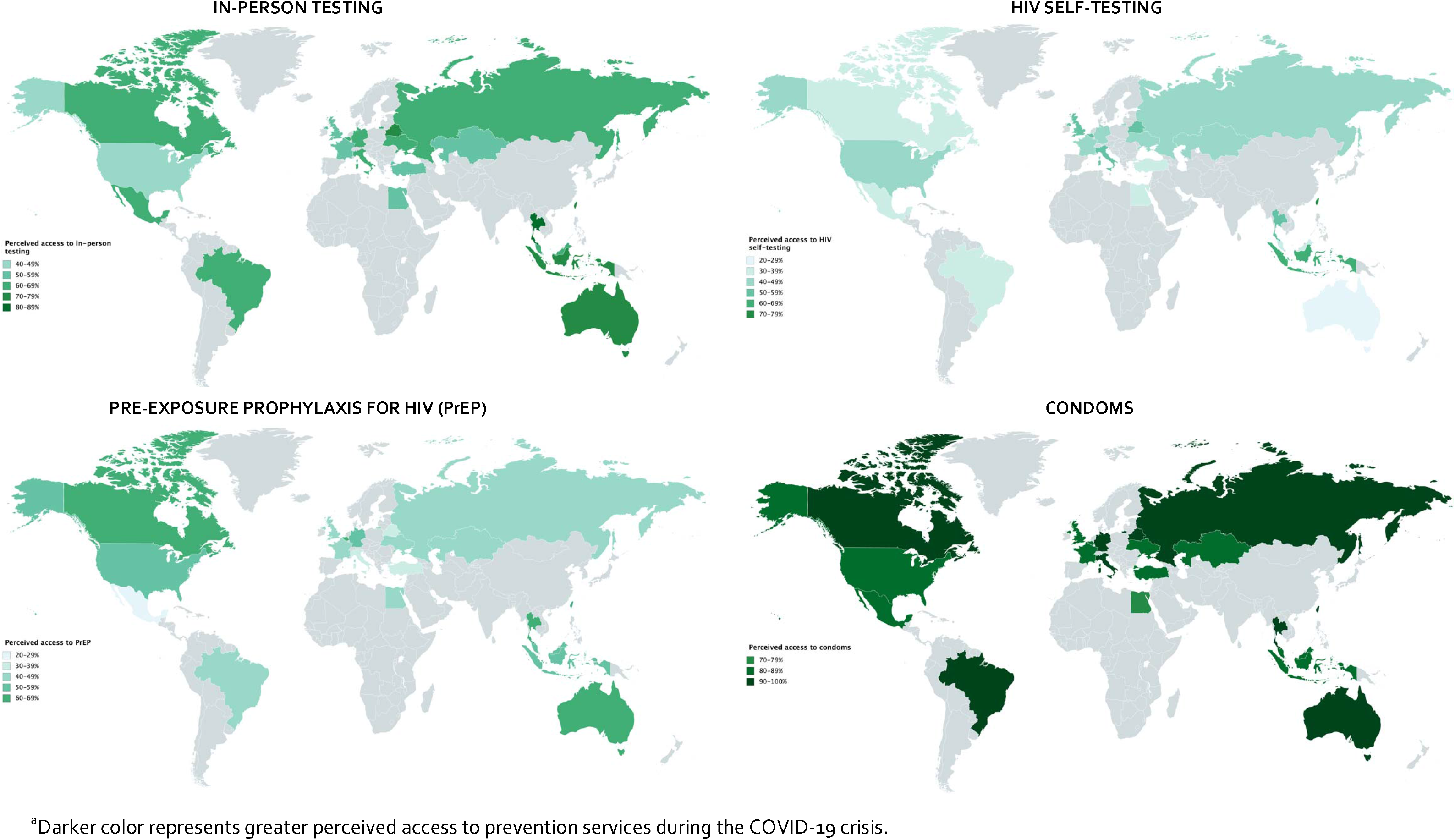
Prevalence of perceived access to HIV prevention services in 20 included countries^a^

### Access to HIV Treatment Services

A total of 1264 individuals, or 12%, reported they were living with HIV, and 94% of them reported being on treatment (n = 1186). Close to one fifth (19.7%) of participants living with HIV reported being unable to access their HIV provider either in-person or via telemedicine (218/1105). About 14% (157/1105) were able to access their provider through innovations in telemedicine, including virtual appointments. Greater than 40% reported being unable to access their provider in Belgium (3/7), Egypt (2/5), and Mexico (28/63). Close to half overall were unable to refill their HIV medicine prescription remotely (n = 562), while 18.6% were able to access their prescription remotely because their provider had made this possible during the COVID-19 pandemic. Over 75% reported being unable to refill their medication remotely in Belarus (11/13), Brazil (127/166), Kazakhstan (13/15), Mexico (60/70), Russia (345/399), and Taiwan (17/21).

### Relationship Between Stringency of Government Response to COVID-19 and Access to HIV Prevention Services

For every 10-point increase in the stringency of the government response to COVID-19, there was a three percent reduction in the prevalence of access to in-person testing (PR: 0.97, 95% CI:[0.96, 0.98]); a six percent reduction in access to HIV self-testing (PR: 0.94, 95% CI:[0.93, 0.95]); a four percent reduction in access to PrEP (PR: 0.96, 95% CI:[0.95, 0.97]); and no significant reduction in access to condoms (PR: 0.99, 95% Cl:[0.99,1.00]). Adjusting for country-level human development index and total health spending per capita, there remained a significant reduction in access to in-person testing (aPR: 0.97, 95% CI:[0.96, 0.98]); access to HIV self-testing (aPR: 0.94, 95% CI:[0.93, 0.95]); and access to PrEP (aPR: 0.95, 95% CI:[0.95, 0.97]) **(Table 2)**.

**Table 2.** Unadjusted and adjusted prevalence ratios for the association of Oxford COVID-19 Government Response Tracker (OxCGRT) Government Response Stringency Index score with access to HIV prevention services among gay and other men who have sex with men participants participating in a global survey implemented via the social media app *Hornet* between April 16^th^ and May 24^th^, 2020^a^.

| Access to... | Crude Prevalence Ratio | 95% Confidence Interval | p-value | Adjusted Prevalence Ratio<sup>b</sup> | 95% Confidence Interval | p-value |
| --- | --- | --- | --- | --- | --- | --- |
| **In-person HIV testing**<br>(n=10396) | 0.97 | 0.96, 0.98 | **<0.001** | 0.97 | 0.96, 0.98 | **<0.001** |
| **HIV self-testing**<br>(n=9335) | 0.94 | 0.93, 0.95 | **<0.001** | 0.94 | 0.93, 0.95 | **<0.001** |
| **Pre-exposure prophylaxis (PrEP)**<br>(n=1892) | 0.96 | 0.95, 0.97 | **<0.001** | 0.95 | 0.95, 0.97 | **<0.001** |
| **Condoms**<br>(n=9542) | 0.99 | 0.99, 1.00 | 0.06 | 0.99 | 0.99, 1.00 | 0.224 |

<sup>a</sup>Given that the exposure was scaled by 10, the interpretation of the prevalence ratio is the relative prevalence of the outcome for every 10-point increase in the Stringency Index. <sup>b</sup>Adjusted for country-level Human Development Index (HDI) and total health spending per capita.

### Sensitivity Analyses

In sensitivity analyses where we varied the dates of the recorded stringency of government response scores, the meaning and interpretation of the results were unchanged **(Appendix)**.

## DISCUSSION

In this global social media sample of over ten thousand MSM from 20 countries, more than half reported a lack of access to HIV self-testing (55%) and PrEP (56%) during the COVID-19 pandemic. About four in ten reported a lack of access to in-person testing, while only about 10% reported a lack of access to condoms. Of the 1264 individuals living with HIV, 20% reported being unable to access their HIV service provider due to the secondary effects of COVID-19 mitigation strategies. Greater stringency of government responses to COVID-19, as measured by the OxCGRT stringency index, was associated with reductions in access to in-person HIV testing, HIV self-testing, and PrEP after accounting for country-level differences in human development and per capita spending on healthcare. These results provide early evidence that, among MSM, efforts to curb the spread of COVID-19 were also associated with disruptions to routine HIV service delivery worldwide.

Globally, governments have implemented physical and social distancing, isolation, and quarantine orders along a range of specificity, as part of a response strategy to community transmission of SARS-CoV-2. While in many places, these strategies have resulted in reduced SARS-CoV-2 incidence, the impact of these mitigation strategies on both perceived and actual access to services for non-COVID-19 competing health issues remains largely unknown. In one study at Beth Israel Deaconess Medical Center in Boston, it was found that patient visits for heart attack and stroke declined by a third or more during the period after government officials declared a state of emergency for COVID-19 compared with the same period in 2019.^26^,^27^ Another study found that there was a reduction in hospital admissions for acute coronary syndrome in England by the end of March, 2020; the authors noted that this reduction likely resulted in preventable deaths and long-term complications of myocardial infarctions.^28^ In a global survey of 155 countries conducted by the World Health Organization, more than half of countries reported partial or complete disruptions in prevention and treatment services for non-communicable diseases, such as cancer, cardiovascular disease, and diabetes.^29^ Lower-income countries were the most affected by these health service disruptions.^29^ A review of the National Syndromic Surveillance Program (NSSP) conducted by the U.S. Centers for Disease Control and Prevention (CDC) found that there were marked declines in emergency departments visits across the United States during the early pandemic period. Others have noted concerns related to how the response to COVID-19 globally will impact vaccine uptake, and subsequently, vaccine-preventable diseases, like measles, pertussis, and rotavirus gastroenteritis.^30^ In a rapid survey conducted among MSM in the United States, men who had sought HIV testing reported challenges in accessing these services due to COVID-19, while a smaller proportion reported difficulty accessing PrEP.^18^

Reduced availability and access to HIV diagnostic, prevention, and treatment services for MSM could lead to increases in new HIV infections, along with a rise in the number of HIV-related deaths. This rise may be particularly problematic in high HIV prevalence settings. In a modeling study of COVID-19 disruptions to the South African context, interruptions to the provision of condoms, PrEP, and ART were predicted to result in tens of thousands of excess deaths due to HIV.^31^ Under conservative model assumptions, predicted excess deaths due to HIV were higher than that from COVID-19.^31^ Existing strategies, such as mobile service delivery, multi-month dispensing, and HIV self-test kits, along with innovative strategies, like app-based dissemination of information to stigmatized populations and community-organized outreach, are needed to overcome the interruptions in access to care created by COVID-19 mitigation efforts.^31^ In this sample of MSM, close to 1 in 5 were able to refill their medication remotely because participants reported that their providers made it possible during COVID-19. In order to avoid further marginalization, sustained efforts to engage community and network leadership in their development may be especially important for MSM.

This study has some limitations. First, the Hornet survey is based on a convenience sample. Those who received the survey had to be active Hornet users and have used the app in the last year, implying that they had access to a smartphone and Wi-Fi or data and knowledge of the app. Those who participated in the survey had to take the time to complete the survey. Sub-Saharan Africa, a region of the world heavily affected by HIV, is not represented in our sample. While these results are very likely not representative of all MSM globally, they provide timely insights into potential disruptions to HIV prevention and treatment services. Second, between 2% and 14% of responses for the primary prevention outcome variables were missing. Complete case analysis was performed and incomplete responses were assumed to be missing completely at random (MCAR) as no discernable patterns of missingness were observed. In the case that data were missing not at random (MNAR), with the missingness due to some other variable not captured in the dataset, our effect estimates may be biased. While this is a limitation of a survey made available through an app-based platform, data were still captured on a large number of individuals and these data provide rapid insights on current access to services. Third, in this analysis we examined country-level stringency of response, but this may only provide part of the picture if there is significant heterogeneity in stringency of the response within a country, for example province, region, or even city. Finally, the primary prevention outcomes that were measured in this survey asked about perceived access during the COVID-19 pandemic, and therefore do not directly capture interruptions in access to care.

During the early period of the pandemic, it was found that men who have sex with men responding to an app-based survey reported low levels of access to HIV services. Moreover, increasingly stringent government responses were associated with decreased access to in-person HIV testing, HIV self-testing and PrEP. Many people living with HIV were not able to access their treatment. To minimize increases in HIV-related morbidity and mortality, innovative strategies are needed to facilitate access to HIV-related diagnostic, prevention, and treatment services during this and potential future waves of COVID-19.

## Data Availability

The data that support the findings of this study are available from Hornet but restrictions apply to the availability of these data, which are not publicly available. Data from the OxCGRT Government Response Stringency Index are publicly available through the Oxford COVID-19 Government Response Tracker online reporting systems.

https://www.bsg.ox.ac.uk/research/research-projects/coronavirus-government-response-tracker

